# Limited-site adaptation reveals representation-specific MRI transportability for lymphovascular invasion in endometrial cancer

**DOI:** 10.64898/2026.07.14.26358100

**Authors:** Daniel A. Di Giovanni, Akiyo Takada, Takuro Horikoshi, Takahiro Tsuboyama, Hajime Yokota, Rita Zakarian, Yuka Matsumoto, Martin Vallières, Caroline Reinhold

**Affiliations:** Department of Diagnostic Radiology, McGill University, Montreal, Quebec, Canada; Augmented Intelligence & Precision Health Laboratory, Research Institute of the McGill University Health Centre, Montreal, Quebec, Canada; Diagnostic Radiology and Radiation Oncology, Chiba University Graduate School of Medicine, Chiba, Japan; Department of Radiology, Chiba University Hospital, Chiba, Japan; Department of Radiology, The University of Osaka Graduate School of Medicine, Osaka, Japan; Research Institute of the McGill University Health Centre, Montreal, Quebec, Canada; Medical Physics Unit, Department of Oncology, McGill University, Montreal, Quebec, Canada

**Keywords:** Endometrial neoplasms, Lymphovascular invasion, Magnetic resonance imaging, Radiomics, Domain adaptation

## Abstract

**Objectives:** To determine how clinically anchored MRI representations behave from supportive internal testing through locked institutional transfer and limited target-site adaptation for prediction of substantial lymphovascular space invasion (LVSI) in endometrial cancer.

**Materials and Methods:** This retrospective two-center study included 206 women undergoing preoperative 3-T MRI between March 2016 and March 2023. Hospital A provided training (n = 117) and supportive internal testing (n = 13); Hospital B provided strict external testing (n = 76; 12 positive). Age and CA125 formed the clinical baseline and were included in every imaging model. In 200 repeated stratified two-fold Hospital B splits, each patient was evaluated outside the adaptation half. Models remained locked; adaptation changed only the threshold or applied no-empirical-Bayes (no-EB) ComBat before threshold selection.

**Results:** DenseNet121, U-NEXtractor, and fusion U-NEXtractor achieved internal AUC 1.000 but strict external AUCs of 0.685, 0.671, and 0.669, respectively. At development-locked thresholds, sensitivities were 0.167, 0.583, and 0.417. Threshold adaptation increased median sensitivity to 0.833 for all three, with specificities of 0.625, 0.500, and 0.500. No-EB ComBat changed median AUC by −0.007, −0.007, and −0.014, respectively, versus strict transfer. U-NEXtractor prediction ranking was most stable (median Spearman ρ, 0.999), whereas fusion did not improve transportability. Empirical-Bayes ComBat lacked complete repeated-split coverage.

**Conclusion:** Strong internal performance did not establish transportability. Limited target-site threshold adaptation recovered operating sensitivity without improving discrimination; feature harmonization produced no consistent AUC gain and affected representations differently.

## Introduction

Substantial lymphovascular space invasion (LVSI) is associated with nodal metastasis, recurrence, and poorer survival in endometrial cancer and now contributes to 2023 International Federation of Gynecology and Obstetrics staging [1]. Because LVSI is usually established only on final hysterectomy histopathology, a reliable preoperative predictor would add prognostic information not otherwise available from routine local staging. Pelvic MRI is the reference standard for local staging, and standard MRI sequences have shown promise for LVSI prediction [2-5].

Prior MRI radiomics and deep-learning studies have reported encouraging discrimination for LVSI [5-8], but clinical translation depends on more than within-site AUC. Radiomic features are sensitive to acquisition and preprocessing, and small-sample model development can exaggerate apparent performance [9-12]. For deployment, models must remain informative when scanners, coils, acquisition planes, and protocols differ between development and target sites.

An unresolved question is whether representation choice influences transportability when acquisition differences are preserved rather than harmonized away. In small, high-dimensional imaging cohorts, classifier flexibility and unstable model selection can compound overfitting [13-16]. Similar discrimination may also yield different operating behavior when calibration or decision thresholds shift across institutions.

This retrospective two-center study therefore compared handcrafted radiomics with seed-pooled deep representations for MRI prediction of substantial LVSI. All preprocessing, model selection, calibration, and a sensitivity-targeted threshold were locked at the development site before evaluation at a second institution with different scanner, coil, acquisition plane, and protocol. The purpose was to determine whether representation choice affected external discrimination and operating-point transportability under cross-site acquisition shift, following principles of rigorous external validation and transparent prediction-model reporting [17, 18].

We extended that locked transfer experiment with a clinical age-plus-CA125 baseline and a leakage-free, repeated cross-fitted adaptation analysis. The prespecified questions were whether limited target-site threshold adaptation could recover operating performance without changing discrimination, whether ComBat feature alignment produced consistent additional benefit, and which imaging representation preserved prediction ranking most reliably across testing conditions.

## Materials and Methods

### Study design, ethics, and patient cohort

Institutional review boards at both participating institutions approved this retrospective two-center secondary analysis and waived written informed consent. The McGill University Health Centre Research Ethics Board, Cells, Tissues, Genetics & Qualitative Research Panel, approved the local study (project MP-37-2023-8859; IRB00010120; initial approval December 23, 2022; current renewal through December 23, 2026). Among 266 initially retrieved patients, 60 were excluded for invisible tumor, insufficient image quality, incomplete MRI sequences, insufficient pathology, an MRI-to-surgery interval greater than 6 weeks in LVSI-positive cases, or preoperative chemotherapy or hormone therapy (Figure 1). The final cohort comprised 206 women with histologically confirmed endometrial cancer who underwent preoperative 3-T pelvic MRI from March 2016 to March 2023. Hospital A (n = 130) was divided by patient into training (n = 117) and supportive internal testing (n = 13; two positive cases). Hospital B (n = 76; 12 positive cases) was the target-site cohort. No formal sample-size calculation was performed; all eligible patients were analyzed. The cohort overlaps a prior radiomics feasibility study [4].

**Figure 1.**
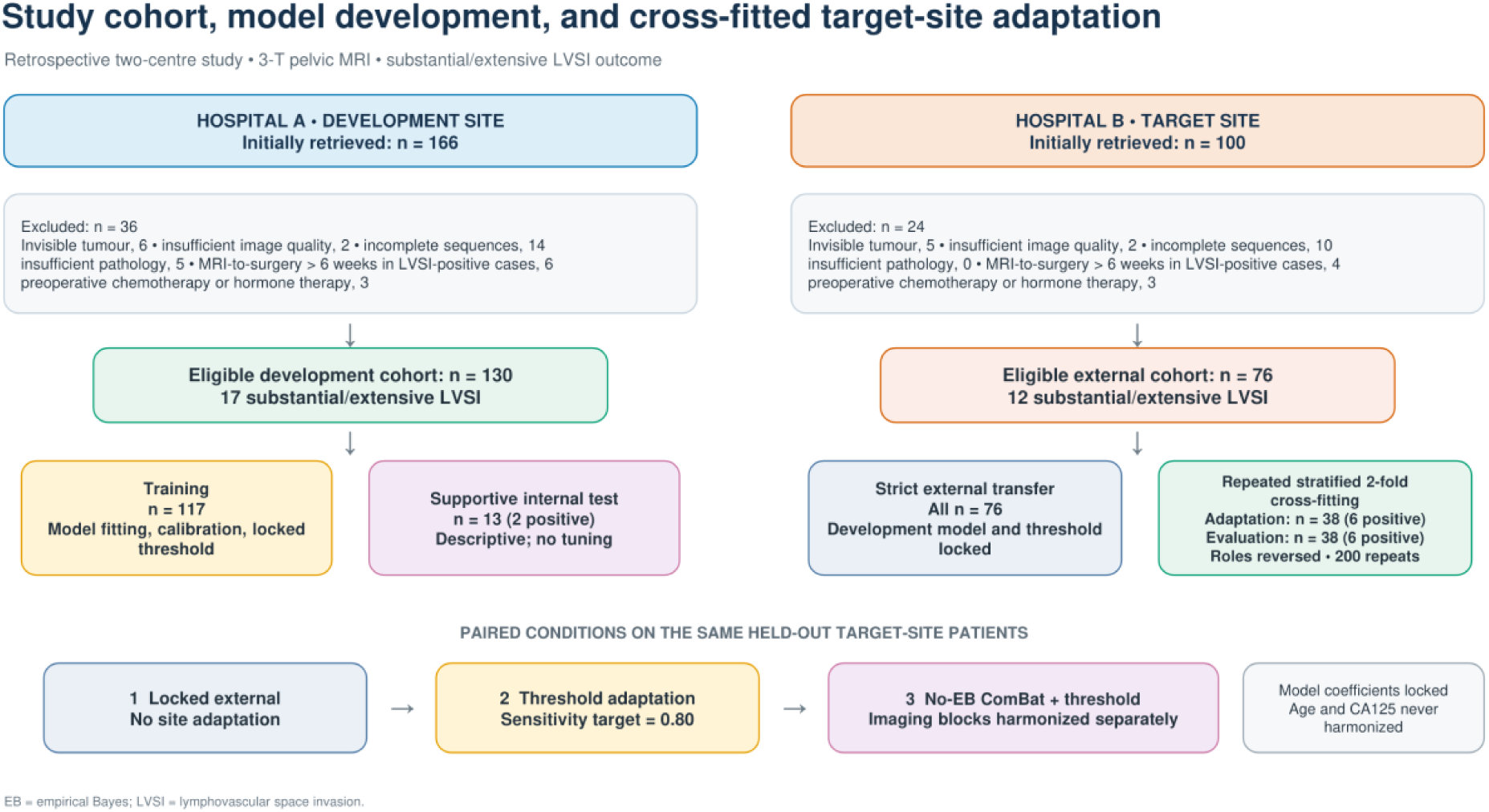
Study cohort, model development, and cross-fitted target-site adaptation. Of 266 initially retrieved patients, 206 were eligible. Hospital A provided training (n = 117) and supportive internal testing (n = 13). Hospital B underwent both strict external transfer and 200 repeated stratified two-fold adaptation/evaluation splits. Each target-site patient was evaluated outside their own adaptation half. Model coefficients remained locked; age and CA125 were included in every model and never harmonized. EB = empirical Bayes; LVSI = lymphovascular space invasion.

### MRI acquisition, segmentation, and outcome

Analyses used T2-weighted imaging, reduced field-of-view diffusion-weighted imaging (FOCUS), and apparent diffusion coefficient (ADC) maps acquired on 3.0-T MR750 or SIGNA Architect systems (GE HealthCare). Scanner model, receiver coil, acquisition plane, field of view, and timing parameters differed by site (Table 2; Supplementary Table S1). A radiologist with 10 years of gynecologic oncology MRI experience manually segmented each tumor in three dimensions; a second radiologist with 20 years of experience repeated segmentations for the reproducibility subset. Readers knew the cancer diagnosis but were blinded to LVSI status. The reference standard was substantial/extensive LVSI on final hysterectomy pathology, modeled against negative/focal LVSI.

### Clinical, radiomic, and deep representations

Age and serum CA125 constituted a separate clinical block. Missing CA125 values were median-imputed within each training fold and then scaled using training-fold parameters. The fitted imputer and scaler were applied unchanged to held-out patients. Age and CA125 were included in every imaging model but were never feature-harmonized.

Radiomic features were extracted separately from T2-weighted images, FOCUS, and ADC maps with an Image Biomarker Standardization Initiative-oriented workflow [12] implemented in PyRadiomics [19] and SimpleITK. Boundary perturbations, fixed-bin-width discretization, isotropic resampling, three-dimensional texture, wavelet, and Laplacian-of-Gaussian features are summarized in the Supplementary Methods; the workflow has been reported previously [4].

Within training data, radiomic features were retained when inter-reader Spearman correlation was at least 0.80, pruned at pairwise absolute correlation greater than 0.95, and screened by Mann–Whitney U testing and repeated elastic-net selection. Features selected in at least 50% of repeats were retained before the clinical block was appended.

Each image-mask pair was cropped around the lesion, resized to 96 × 96 × 96 voxels, and z-score normalized; ADC values were constrained to nonnegative values. Three-dimensional ResNet18 [20], DenseNet121 [21], and U-NEXtractor-style [22] backbones were initialized randomly and trained from scratch. ResNet18 and DenseNet121 could receive image and mask channels; U-NEXtractor used image-only input with classification and auxiliary segmentation heads. Penultimate-layer embeddings were exported.

Three seeds (42, 43, and 44) were trained per backbone within the 117-patient training cohort. Each seed used a stratified 80:20 fit/validation split, AdamW optimization (learning rate, 3 × 10−5; weight decay, 1 × 10−4), up to 50 epochs, 10-epoch early stopping, and validation PR AUC for checkpoint selection. Penultimate-layer embeddings were averaged elementwise across seeds. Clinical, radiomics-plus-clinical, deep-plus-clinical, and radiomics-plus-deep-plus-clinical fusion views were matched by patient identifier.

### Locked development pipeline

Within each development fold, radiomics were robust-scaled; deep embeddings were standardized and reduced to 24 principal components. Fusion blocks were processed separately before concatenation. Elastic-net logistic regression was the prespecified downstream classifier. Repeated out-of-fold training probabilities were Platt-calibrated, and a bootstrap-stabilized threshold targeting sensitivity of 0.80 was selected entirely from development data. Preprocessing, selected features, principal-component transformations, classifier coefficients, calibrator, and threshold were then locked. The internal and strict external cohorts contributed no information to model fitting or selection.

### Target-site adaptation experiment

Hospital B was analyzed with 200 repeated stratified two-fold splits (seed 20260824). In each direction, 38 patients, including six positive cases, formed a labeled adaptation half and the other 38, also including six positive cases, formed an untouched evaluation half; roles were then reversed. Thus, every Hospital B patient was evaluated once per repeat without contributing to their own adaptation, and paired strict-versus-adapted results used exactly the same held-out patients.

The primary adaptation changed only the decision threshold, selecting the value in the adaptation half that targeted sensitivity of 0.80. Feature harmonization used neuroHarmonize 2.5.1 with Hospital A as the explicit reference batch. Radiomic and deep blocks were harmonized separately, with source-scaled age and CA125 included as covariates but not transformed. Empirical-Bayes and no-empirical-Bayes (no-EB) ComBat were evaluated separately [29–31]. Model coefficients were never refitted. Automated safety checks rejected transformations with RuntimeWarnings, material source-reference changes, or extreme target-source feature values; conditions without complete repeated-split coverage were excluded from performance interpretation.

### Performance, stability, and statistical analysis

Discrimination was summarized by receiver operating characteristic AUC and PR AUC. Locked operating behavior was summarized by sensitivity, specificity, balanced accuracy, and confusion-matrix counts. Strict external AUCs received class-stratified patient-level bootstrap 95% confidence intervals (5,000 resamples). Paired class-stratified bootstrap comparisons quantified strict external AUC and PR AUC differences versus the clinical baseline. Continuous cohort variables were compared with Mann–Whitney U tests and categorical variables with Fisher exact or chi-square tests, as appropriate. No multiplicity adjustment was applied; two-sided p < 0.05 indicated statistical significance.

Adaptation results are medians across 200 repeats with 2.5th–97.5th percentile repeated-split instability intervals, not confidence intervals or independent validations. Paired changes were calculated within repeat against strict predictions on the same held-out patients. Prediction stability under harmonization was summarized by Spearman rank correlation, mean absolute probability change, and locked-classification agreement. Feature shift was summarized by standardized mean differences and model-importance-weighted absolute SMD. Reporting followed TRIPOD+AI and CLAIM guidance [18, 32].

Analyses used Python 3.11; PyRadiomics 2.2; SimpleITK 2.5; PyTorch 2.12; MONAI 1.5; scikit-learn 1.7; and neuroHarmonize 2.5.1.

## Results

The final cohort comprised 206 of 266 initially retrieved women: 130 at Hospital A (17 substantial/extensive LVSI-positive) and 76 at Hospital B (12 positive) (Figure 1). Hospital A exclusions totaled 36 and Hospital B exclusions totaled 24. Mean age was 60.3 ± 12.6 years at Hospital A and 59.0 ± 10.2 years at Hospital B; LVSI prevalence, histologic type, CA125, and FIGO stage did not differ significantly between sites (all p ≥ 0.08) (Table 1).

**Table 1.** Cohort characteristics and analysis populations.

| Characteristic | Hospital A | Hospital B | p value |
| --- | --- | --- | --- |
| Number of patients | 130 | 76 |  |
| Age, mean ± SD (years) | 60.3 ± 12.6 | 59.0 ± 10.2 | 0.76 |
| LVSI |  |  | 1.00 |
| Negative/focal | 113 | 64 |  |
| Substantial/extensive | 17 | 12 |  |
| Histologic type |  |  | 0.86 |
| Type 1 (low risk) | 104 | 61 |  |
| Type 2 (high risk) | 26 | 15 |  |
| CA125, mean ± SD | 57.8 ± 187.5 | 26.6 ± 39.3 | 0.14 |
| FIGO stage |  |  | 0.08 |
| I | 89 | 60 |  |
| II | 15 | 2 |  |
| III | 25 | 12 |  |
| IV | 1 | 2 |  |

CA125 = cancer antigen 125; FIGO = International Federation of Gynecology and Obstetrics; LVSI = lymphovascular space invasion; SD = standard deviation. FIGO stage reflects the stage recorded in the source clinical record.

**Table 2.** Site-specific MRI acquisition differences relevant to institutional transfer.

| Parameter | Hospital A | Hospital B |
| --- | --- | --- |
| Scanner and coil | 3.0-T GE MR750; 32-channel Body Array coil | 3.0-T GE SIGNA Architect; 30-channel Adaptive Imaging Receive coil |
| T2-weighted imaging | Axial oblique; TR/TE 7200/100 ms; FOV 220 × 220 mm; matrix 512 × 512; slice/spacing 4/4.8 mm | Axial oblique; TR/TE 4100/80 ms; FOV 288 × 288 mm; matrix 512 × 512; slice/spacing 4/4 mm |
| rFOV DWI / FOCUS | Axial oblique; TR/TE 3000/60 ms; FOV 200 × 200 mm; matrix 256 × 256; slice/spacing 4/4.8 mm; b = 1000 s/mm<sup>2</sup> | Sagittal; TR/TE 4500/65 ms; FOV 110 × 110 mm; matrix 256 × 256; slice/spacing 4/4 mm; b = 1000 s/mm<sup>2</sup> |

DWI = diffusion-weighted imaging; FOCUS = reduced field-of-view diffusion-weighted imaging; FOV = field of view; rFOV = reduced field of view; TE = echo time; TR = repetition time.

### Supportive internal testing and strict external transfer

Internal AUCs were 0.727 for the clinical baseline, 0.682 for radiomics plus clinical, and 1.000 for DenseNet121 plus clinical, U-NEXtractor plus clinical, and fusion U-NEXtractor plus clinical. Because the internal cohort contained only 13 patients and two positive cases, these values are descriptive rather than precise estimates. Internal balanced accuracy was 0.545, 0.614, 0.955, 0.818, and 0.864, respectively (Table 3).

**Table 3.**
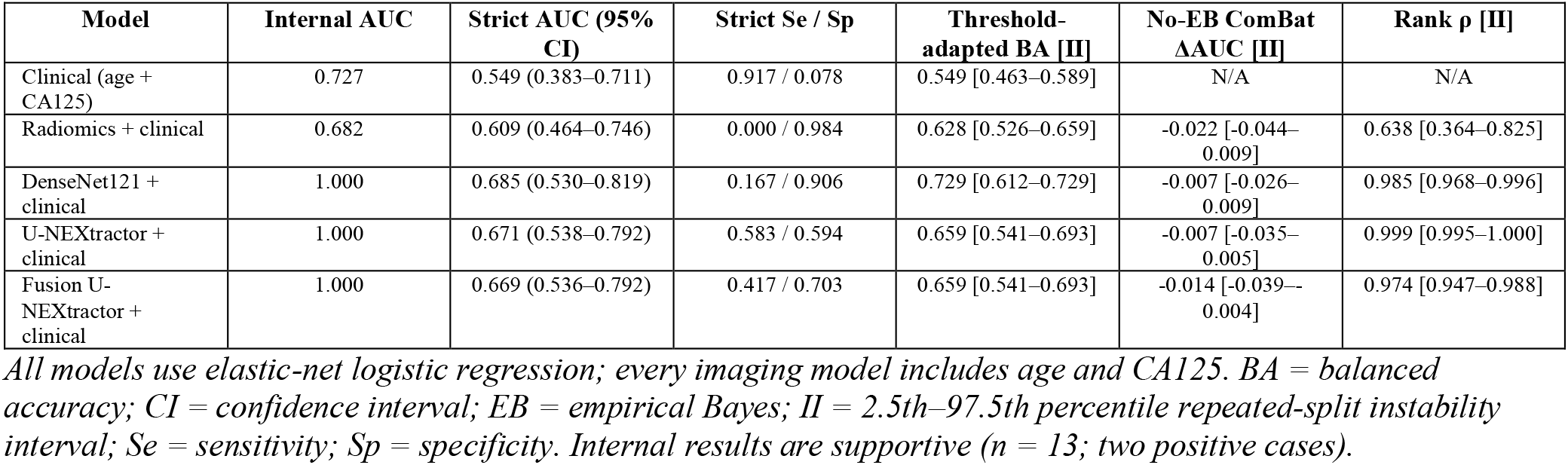
Primary model performance across testing and adaptation conditions.

Under strict external transfer, AUCs were 0.549 (95% CI, 0.383–0.711) for the clinical baseline, 0.609 (0.464–0.746) for radiomics plus clinical, 0.685 (0.530–0.819) for DenseNet121 plus clinical, 0.671 (0.538–0.792) for U-NEXtractor plus clinical, and 0.669 (0.536–0.792) for fusion U-NEXtractor plus clinical. None of the imaging-model AUC differences versus the clinical baseline reached statistical significance (all p ≥ 0.103; Supplementary Table S8).

Development-locked thresholds exposed different failure patterns. The clinical baseline detected 11 of 12 positive cases but misclassified 59 of 64 negative cases (sensitivity, 0.917; specificity, 0.078). Radiomics plus clinical detected none of 12 positive cases while correctly classifying 63 of 64 negative cases. DenseNet121 plus clinical detected 2 of 12 positive cases (sensitivity, 0.167; specificity, 0.906); U-NEXtractor plus clinical detected 7 of 12 (0.583; 0.594); and fusion U-NEXtractor plus clinical detected 5 of 12 (0.417; 0.703).

### Threshold adaptation

Threshold adaptation left patient ranking and AUC unchanged by design but shifted operating performance (Figures 2 and 3). Median sensitivity/specificity became 0.750/0.344 for the clinical baseline, 0.750/0.469 for radiomics plus clinical, 0.833/0.625 for DenseNet121 plus clinical, and 0.833/0.500 for both U-NEXtractor plus clinical and fusion U-NEXtractor plus clinical. Median balanced-accuracy gains versus strict transfer were 0.052, 0.135, 0.193, 0.070, and 0.099, respectively; their instability intervals crossed zero for the clinical, U-NEXtractor, and fusion U-NEXtractor models (Figure 3).

**Figure 2.**
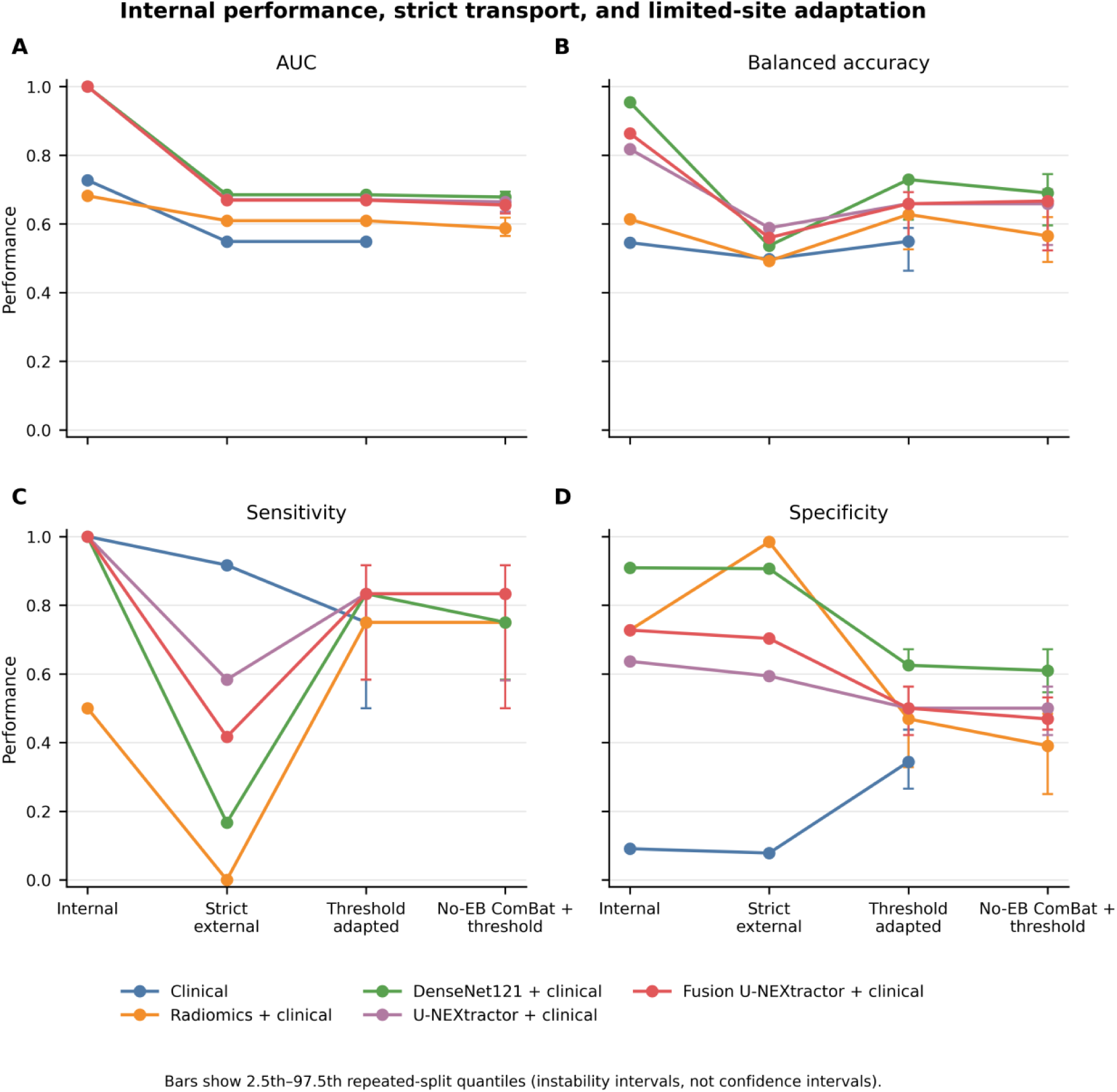
Internal performance, strict transfer, and limited-site adaptation. Panels show AUC (A), balanced accuracy (B), sensitivity (C), and specificity (D) for five prespecified clinically anchored models. Internal and strict values are fixed estimates; adapted values are medians across 200 repeated stratified two-fold splits. No-EB ComBat was applied to imaging blocks before threshold adaptation. Error bars show 2.5th–97.5th percentile repeated-split instability intervals, not confidence intervals. AUC = area under the receiver operating characteristic curve; EB = empirical Bayes.

**Figure 3.**
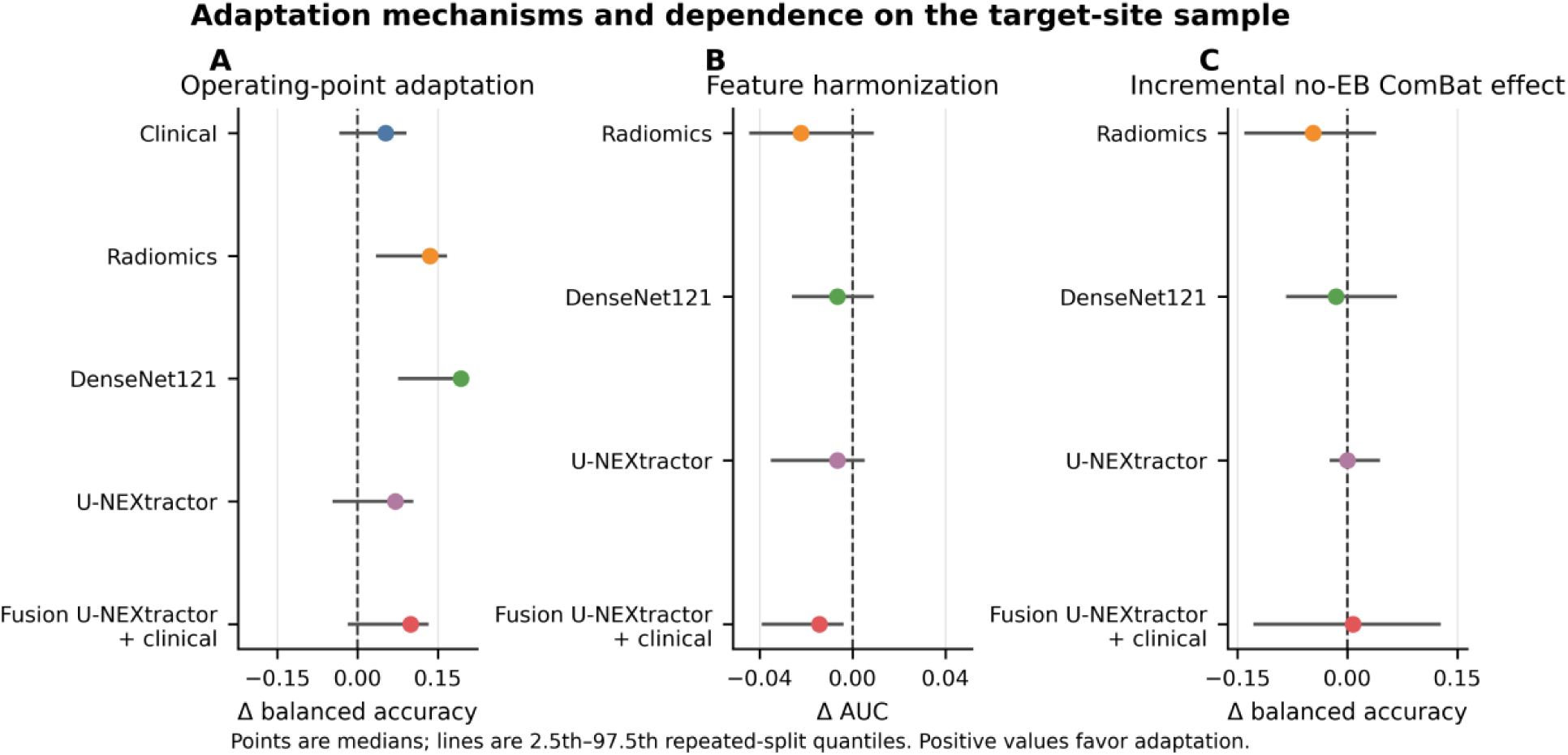
Adaptation mechanisms and dependence on the target-site sample. Panel A shows the balanced-accuracy change after threshold adaptation versus strict transfer. Panel B shows the AUC change after no-EB ComBat versus strict transfer. Panel C shows the incremental balanced-accuracy change when no-EB ComBat preceded threshold adaptation versus threshold adaptation alone. Points are medians; lines are 2.5th–97.5th percentile repeated-split instability intervals. Positive values favour adaptation.

### Feature harmonization and representation stability

No-EB ComBat achieved complete 200-repeat coverage for all seven imaging models. Relative to strict transfer, median AUC changes were −0.022 for radiomics plus clinical, −0.007 for DenseNet121 plus clinical, −0.007 for U-NEXtractor plus clinical, and −0.014 for fusion U-NEXtractor plus clinical. Adding no-EB ComBat before threshold adaptation did not produce a consistent balanced-accuracy advantage over threshold adaptation alone: median incremental changes were −0.047, −0.016, 0.000, and 0.008, respectively (Figure 3; Supplementary Tables S4 and S5).

Prediction perturbation differed by representation (Figure 4). Median strict-versus-harmonized rank correlation was 0.999 for U-NEXtractor plus clinical, 0.985 for DenseNet121 plus clinical, 0.974 for fusion U-NEXtractor plus clinical, and 0.638 for radiomics plus clinical. Median mean absolute probability change was 0.010, 0.039, 0.020, and 0.160, respectively. Although no-EB ComBat reduced the importance-weighted absolute SMD from 0.812 to 0.162 for radiomics and from 0.257 to 0.161 for DenseNet121, it increased that summary from 0.063 to 0.159 for U-NEXtractor while its prediction ranking remained nearly unchanged. Empirical-Bayes ComBat had zero complete repeats for radiomics and fusion models and only 7.0%–12.5% complete-repeat coverage for deep-representation models because divide-by-zero runtime warnings triggered the safety gate; it was therefore retained only as a diagnostic analysis (Supplementary Figure S3).

**Figure 4.**
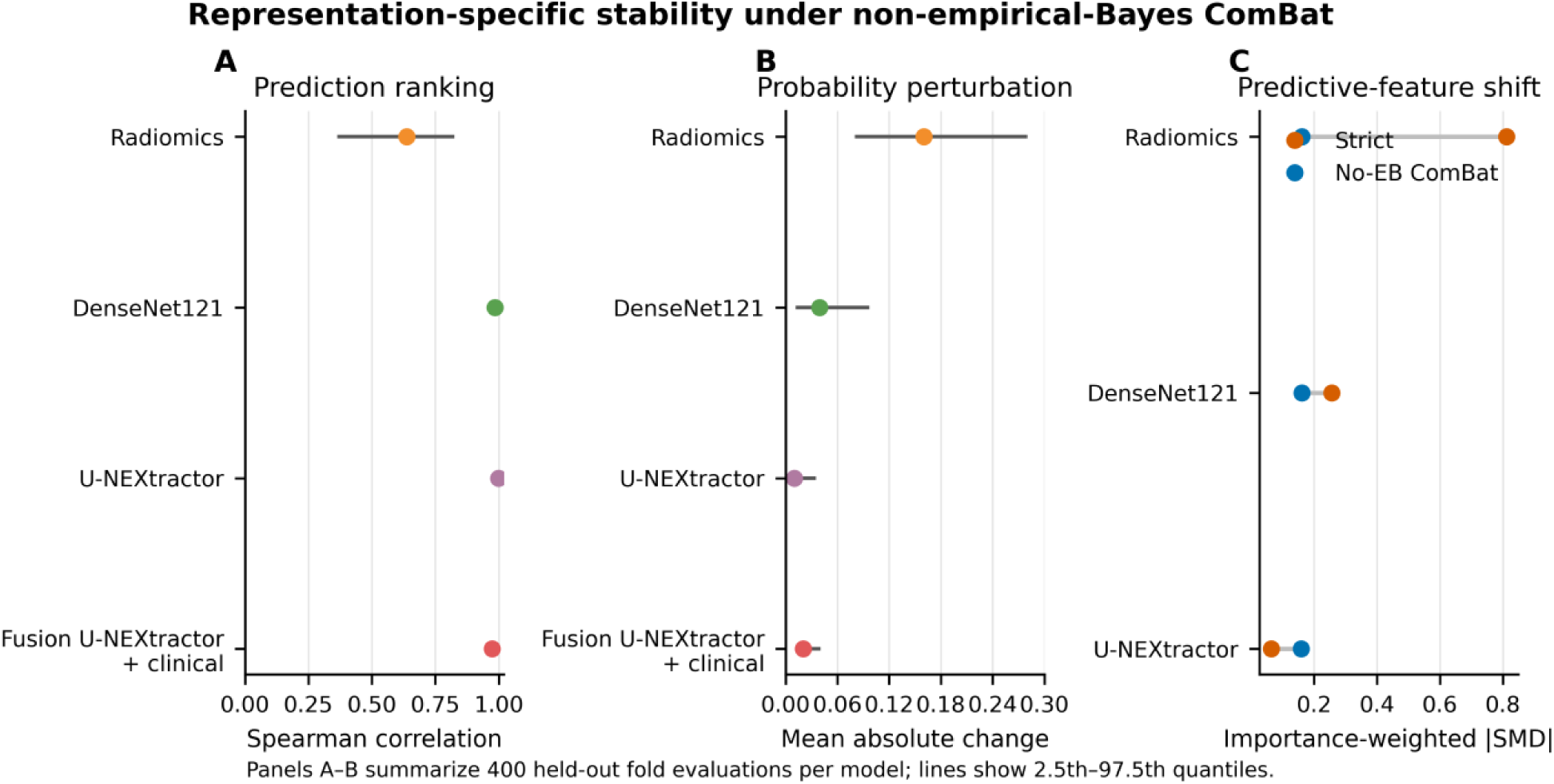
Representation-specific stability under no-empirical-Bayes ComBat. Panel A shows Spearman correlation between strict and harmonized patient ranking. Panel B shows mean absolute probability change. Panel C shows the median model-importance-weighted absolute standardized mean difference before and after harmonization. Panels A and B summarize 400 held-out-fold evaluations per model; lines show 2.5th–97.5th percentile repeated-split instability intervals. CA125 = cancer antigen 125; SMD = standardized mean difference.

## Discussion

This clinically anchored, two-center study shows that apparently excellent internal MRI model performance did not establish institutional transportability. The main distinction was mechanistic: limited target-site threshold adaptation recovered sensitivity without improving discrimination, whereas no-EB ComBat produced no consistent AUC gain and substantially altered less stable representations. U-NEXtractor predictions preserved ranking most reliably across repeated target-site samples, and adding radiomics in fusion did not provide a robust transport benefit.

Prior LVSI studies have reported higher AUCs for clinical-radiomics and integrated radiomics-deep-learning models [5-8]. The present study addressed a different question, namely, performance after model locking when scanner and protocol differences were deliberately preserved between institutions. This design produced more modest headline AUCs but directly tested whether MRI-derived representations and operating thresholds remained transportable across sites.

The comparison with an age-plus-CA125 baseline is central. The clinical model was highly sensitive but clinically impractical at its locked threshold because it overcalled 59 of 64 external negative cases. Radiomics plus clinical showed the opposite failure, missing every positive case. Deep representations occupied different points between these extremes. These error profiles demonstrate why a single AUC, even when externally estimated, cannot describe whether a transferred model has a usable operating point.

Threshold adaptation is not model improvement in the discrimination sense. It uses labeled target-site cases to choose an operating point and therefore changes sensitivity and specificity while leaving AUC unchanged. Cross-fitting prevented an evaluated patient from contributing to their own threshold, but the resulting repeated-split intervals describe dependence on the small adaptation sample rather than a new external-validation confidence interval. The DenseNet121 model showed the largest median balanced-accuracy recovery, yet its interval remained wide, underscoring the uncertainty of six-positive adaptation halves.

ComBat findings require equally careful interpretation. The method was developed for batch-effect adjustment, and reference-site implementations can reduce measured feature differences [29–31]. Here, no-EB ComBat reduced radiomic feature shift but did not improve radiomic AUC, whereas U-NEXtractor ranking remained stable even when the importance-weighted feature-shift summary increased. Thus, distributional alignment and predictive stability were not interchangeable. Empirical-Bayes estimation was numerically unreliable in these small adaptation halves; presenting only successful splits would have overstated apparent performance, so incomplete conditions were excluded.

Seed pooling was a secondary design choice rather than the primary source of novelty. U-NEXtractor showed substantial seed-to-seed variability, and averaging penultimate embeddings across three initializations provided a single representation for downstream testing; this approach is consistent with evidence that training randomness can materially affect deep medical-imaging models [23].

These results reinforce why external validation of imaging prediction models should report calibration, threshold-based measures, and error counts alongside discrimination. A model may retain ranking information while a decision threshold selected at the development site fails after transfer. Calibration, risk distributions, and prespecified operating thresholds therefore provide complementary information to AUC [24-28].

The study has limitations. The internal cohort contained only 13 patients and two positive cases, making its perfect deep-model AUCs supportive rather than definitive. Hospital B contained 12 positive cases; repeated cross-fitting uses those same 76 patients and is not an independent third validation cohort. Both sites used 3-T systems from one vendor, limiting the range of acquisition shift. The retrospective cohort and manual segmentations may introduce selection and measurement bias. The adaptation analysis used labeled target-site outcomes and should not be interpreted as unsupervised deployment. Multiple representations and adaptation mechanisms were evaluated without multiplicity adjustment. Finally, no human-reader comparison was performed, and none of the imaging models significantly exceeded the clinical baseline in external AUC.

In conclusion, institutional transfer exposed representation-specific failure that internal AUC concealed. Limited target-site threshold adaptation could recover operating sensitivity, but feature harmonization did not reliably improve discrimination and was sensitive to both representation and estimation method. U-NEXtractor preserved prediction ranking most consistently, while fusion offered no robust advantage. Larger multicenter studies should validate whether stability across adaptation samples predicts real-world deployment performance.

## Data Availability

All data produced in the present study are available upon reasonable request to the authors

## Abbreviations

ADC: apparent diffusion coefficient
AUC: area under the receiver operating characteristic curve
CA125: cancer antigen 125
EB: empirical Bayes
LVSI: lymphovascular space invasion
PR AUC: area under the precision-recall curve
rFOV DWI: reduced field-of-view diffusion-weighted imaging
SMD: standardized mean difference.

## Declarations

### Ethics approval and consent to participate

Institutional review boards at both participating institutions approved this retrospective two-center secondary analysis and waived written informed consent. The McGill University Health Centre Research Ethics Board, Cells, Tissues, Genetics & Qualitative Research Panel, approved the local study under project MP-37-2023-8859 (IRB00010120); initial approval was December 23, 2022, with current renewal through December 23, 2026.

### Funding

This work received no specific funding.

### Competing interests

The authors declare no competing interests.

### Data and code availability

Deidentified data and analysis code may be made available by the corresponding author upon reasonable request, subject to institutional approvals and data-use restrictions.

## Acknowledgements

OpenAI ChatGPT/Codex was used for language editing, code-assisted document preparation, and consistency checks. The authors reviewed all outputs and take responsibility for the final content.

## Notes

### Competing Interest Statement

The authors have declared no competing interest.

### Author Declarations

The institutional review board of McGill University approved this retrospective study and waived the requirement for informed consent. All analyses used coded, deidentified study data.

### Summary of Updates

This revision materially expands the analysis and reframes the study around clinically anchored model transportability and limited target-site adaptation. The patient cohort, MRI examinations, and substantial/extensive lymphovascular space invasion outcome are unchanged. The title and abstract were revised to reflect the expanded study question. Age and CA125 are now presented as a clinical baseline and are included consistently in every imaging model. The Methods now describe supportive internal testing, strict locked external transfer, and 200 repeated stratified two-fold target-site adaptation experiments in which each Hospital B patient is evaluated outside their own adaptation half. Threshold adaptation, intercept-only recalibration, and reference-site ComBat harmonization are separated analytically. The Results now compare internal, strict external, and adapted external performance; quantify representation-specific prediction stability; and report safety-gated empirical-Bayes and no-empirical-Bayes ComBat analyses. The revised conclusions distinguish recovery of an operating threshold from improvement in discrimination. They also clarify that no imaging model significantly outperformed the clinical baseline in external AUC, U-NEXtractor preserved ranking most consistently, and fusion provided no robust transportability advantage. Figures, Tables, Supplementary Methods, Supplementary Tables, and Supplementary Figures were updated or added accordingly. The reference list was expanded.

