## Supplementary Materials for "Limited-site adaptation reveals representation-specific MRI transportability for lymphovascular invasion in endometrial cancer"

Daniel A. Di Giovanni et al.

### Supplementary Methods

#### Cohort retrieval and partitioning

Hospital A initially contained 166 patients. Exclusions were invisible tumor (n = 6), insufficient image quality (n = 2), incomplete MRI sequences (n = 14), insufficient pathology (n = 5), an MRI-to-surgery interval greater than 6 weeks in LVSI-positive cases (n = 6), and preoperative chemotherapy or hormone therapy (n = 3), leaving 130 patients. Hospital B initially contained 100 patients; corresponding exclusions were 5, 2, 10, 0, 4, and 3, leaving 76 patients. Hospital A was divided into training (n = 117) and supportive internal testing (n = 13). Hospital B underwent both strict external testing and repeated cross-fitted adaptation.

#### Radiomic feature extraction

Radiomic extraction was performed separately for T2-weighted imaging, FOCUS, and ADC maps using PyRadiomics and SimpleITK. Three-dimensional first-order, shape, texture, wavelet, and Laplacian-of-Gaussian features were extracted from original, eroded, and dilated tumor masks to assess boundary sensitivity. Fixed-bin-width discretization and isotropic resampling settings were selected using Hospital A development data only. Reproducibility filtering and all subsequent feature selection were likewise confined to development data.

#### Model specification

The clinical baseline contained age and CA125. Every imaging model contained the identical clinical block: selected radiomics plus clinical, pooled ResNet18/DenseNet121/U-NEXtractor embeddings plus clinical, or selected radiomics plus the corresponding deep embedding plus clinical. Radiomics were robust-scaled. Deep embeddings were standardized and reduced to 24 principal components. Fusion blocks were processed separately before concatenation. Elastic-net logistic regression, out-of-fold Platt calibration, and the development sensitivity-targeted threshold were identical across views.

#### Repeated target-site adaptation

Two hundred repeated stratified two-fold Hospital B splits used seed 20260824. Each adaptation and evaluation half contained 38 patients, including six substantial/extensive LVSI-positive cases. Fold roles were reversed, so all 76 patients were evaluated once per repeat. Primary threshold adaptation targeted sensitivity of 0.80 in the adaptation half. Strict, threshold-adapted, harmonization-only, and harmonization-plus-threshold conditions were paired within the same held-out patients.

#### ComBat implementation and safety gates

neuroHarmonize 2.5.1 used Hospital A as an explicit reference batch. Radiomic and deep blocks were harmonized separately. Source-scaled age and CA125 were included as covariates, but the clinical features themselves were never transformed. Empirical-Bayes (EB) and no-empirical-Bayes (no-EB) modes were evaluated independently. The model classifier and source preprocessing were locked. Each transform had to preserve Hospital A reference features, avoid RuntimeWarnings, retain finite values, and remain within prespecified extreme-shift limits. A model-condition entered performance figures only with complete 200-repeat coverage.

#### Statistical interpretation

Repeated-split 2.5th–97.5th percentile ranges quantify sensitivity to the labeled target-site adaptation sample. They are instability intervals, not confidence intervals, and the 200 repeats are not independent validation cohorts. Class-stratified bootstrap confidence intervals for strict external AUC used 5,000 patient-level resamples. Paired imaging-minus-clinical AUC and PR AUC comparisons used the same class-stratified bootstrap indices.

### Supplementary Tables

**Supplementary Table S1.** Site-specific MRI acquisition parameters.

| Sequence/parameter | Hospital A | Hospital B |
|---|---|---|
| Scanner | 3.0-T GE MR750 | 3.0-T GE SIGNA Architect |
| Coil | 32-channel Body Array | 30-channel Adaptive Imaging Receive |
| T2WI plane / acquisition | Axial oblique / 2D | Axial oblique / 2D |
| T2WI TR/TE (ms) | 7200/100 | 4100/80 |
| T2WI FOV / matrix | 220 × 220 mm / 512 × 512 | 288 × 288 mm / 512 × 512 |
| T2WI slice / spacing | 4 / 4.8 mm | 4 / 4 mm |
| rFOV DWI / FOCUS plane | Axial oblique | Sagittal |
| rFOV DWI TR/TE (ms) | 3000/60 | 4500/65 |
| rFOV DWI FOV / matrix | 200 × 200 mm / 256 × 256 | 110 × 110 mm / 256 × 256 |
| rFOV DWI slice / spacing | 4 / 4.8 mm | 4 / 4 mm |
| High b value | 1000 s/mm² | 1000 s/mm² |
| ADC map | Scanner-generated | Scanner-generated |

**Supplementary Table S2.** Fixed supportive internal and strict external performance for all prespecified models.

| Model | Condition | AUC | PR AUC | Se | Sp | BA | TP | FP | TN | FN |
|---|---|---|---|---|---|---|---|---|---|---|
| Clinical (age + CA125) | Internal | 0.727 | 0.333 | 1.000 | 0.091 | 0.545 | 2 | 10 | 1 | 0 |
| Clinical (age + CA125) | Strict external | 0.549 | 0.197 | 0.917 | 0.078 | 0.497 | 11 | 59 | 5 | 1 |
| Radiomics + clinical | Internal | 0.682 | 0.375 | 0.500 | 0.727 | 0.614 | 1 | 3 | 8 | 1 |
| Radiomics + clinical | Strict external | 0.609 | 0.200 | 0.000 | 0.984 | 0.492 | 0 | 1 | 63 | 12 |
| DenseNet121 + clinical | Internal | 1.000 | 1.000 | 1.000 | 0.909 | 0.955 | 2 | 1 | 10 | 0 |
| DenseNet121 + clinical | Strict external | 0.685 | 0.252 | 0.167 | 0.906 | 0.536 | 2 | 6 | 58 | 10 |
| ResNet18 + clinical | Internal | 0.909 | 0.583 | 0.000 | 0.909 | 0.455 | 0 | 1 | 10 | 2 |
| ResNet18 + clinical | Strict external | 0.612 | 0.196 | 0.000 | 0.938 | 0.469 | 0 | 4 | 60 | 12 |
| U-NEXtractor + clinical | Internal | 1.000 | 1.000 | 1.000 | 0.636 | 0.818 | 2 | 4 | 7 | 0 |
| U-NEXtractor + clinical | Strict external | 0.671 | 0.228 | 0.583 | 0.594 | 0.589 | 7 | 26 | 38 | 5 |

| Model | Condition | AUC | PR AUC | Se | Sp | BA | TP | FP | TN | FN |
|---|---|---|---|---|---|---|---|---|---|---|
| Fusion DenseNet121 + clinical | Internal | 1.000 | 1.000 | 1.000 | 0.727 | 0.864 | 2 | 3 | 8 | 0 |
| Fusion DenseNet121 + clinical | Strict external | 0.569 | 0.190 | 0.250 | 0.656 | 0.453 | 3 | 22 | 42 | 9 |
| Fusion ResNet18 + clinical | Internal | 0.955 | 0.833 | 0.500 | 1.000 | 0.750 | 1 | 0 | 11 | 1 |
| Fusion ResNet18 + clinical | Strict external | 0.568 | 0.184 | 0.000 | 0.891 | 0.445 | 0 | 7 | 57 | 12 |
| Fusion U-NEXtractor + clinical | Internal | 1.000 | 1.000 | 1.000 | 0.727 | 0.864 | 2 | 3 | 8 | 0 |
| Fusion U-NEXtractor + clinical | Strict external | 0.669 | 0.227 | 0.417 | 0.703 | 0.560 | 5 | 19 | 45 | 7 |

**Supplementary Table S3.** Threshold-adapted performance across 200 repeated target-site splits.

| Model | AUC [II] | Sensitivity [II] | Specificity [II] | Balanced accuracy [II] |
|---|---|---|---|---|
| Clinical (age + CA125) | 0.549 [0.549–0.549] | 0.750 [0.500–0.833] | 0.344 [0.266–0.438] | 0.549 [0.463–0.589] |
| Radiomics + clinical | 0.609 [0.609–0.609] | 0.750 [0.583–0.833] | 0.469 [0.328–0.562] | 0.628 [0.526–0.659] |
| DenseNet121 + clinical | 0.685 [0.685–0.685] | 0.833 [0.583–0.833] | 0.625 [0.625–0.672] | 0.729 [0.612–0.729] |
| ResNet18 + clinical | 0.612 [0.612–0.612] | 0.750 [0.583–0.833] | 0.438 [0.328–0.531] | 0.589 [0.510–0.643] |
| U-NEXtractor + clinical | 0.671 [0.671–0.671] | 0.833 [0.583–0.917] | 0.500 [0.422–0.563] | 0.659 [0.541–0.693] |
| Fusion DenseNet121 + clinical | 0.569 [0.569–0.569] | 0.750 [0.500–0.833] | 0.484 [0.250–0.547] | 0.581 [0.466–0.633] |
| Fusion ResNet18 + clinical | 0.568 [0.568–0.568] | 0.750 [0.581–0.833] | 0.391 [0.281–0.438] | 0.570 [0.479–0.620] |
| Fusion U-NEXtractor + clinical | 0.669 [0.669–0.669] | 0.833 [0.583–0.917] | 0.500 [0.422–0.563] | 0.659 [0.541–0.693] |

**Supplementary Table S4.** No-empirical-Bayes ComBat plus threshold-adapted performance.

| Model | AUC [II] | Sensitivity [II] | Specificity [II] | Balanced accuracy [II] |
|---|---|---|---|---|
| Radiomics + clinical | 0.587 [0.565–0.618] | 0.750 [0.581–0.833] | 0.391 [0.250–0.484] | 0.565 [0.490–0.620] |
| DenseNet121 + clinical | 0.678 [0.659–0.694] | 0.750 [0.583–0.833] | 0.609 [0.546–0.672] | 0.690 [0.596–0.745] |
| ResNet18 + clinical | 0.608 [0.586–0.622] | 0.750 [0.581–0.833] | 0.438 [0.328–0.531] | 0.589 [0.508–0.636] |
| U-NEXtractor + clinical | 0.664 [0.635–0.676] | 0.833 [0.581–0.917] | 0.500 [0.422–0.563] | 0.659 [0.539–0.701] |
| Fusion DenseNet121 + clinical | 0.553 [0.527–0.579] | 0.750 [0.500–0.917] | 0.266 [0.125–0.453] | 0.508 [0.461–0.560] |
| Fusion ResNet18 + clinical | 0.499 [0.480–0.516] | 0.750 [0.583–1.000] | 0.219 [0.141–0.297] | 0.500 [0.417–0.586] |
| Fusion U-NEXtractor + clinical | 0.655 [0.630–0.665] | 0.833 [0.500–0.917] | 0.469 [0.438–0.531] | 0.667 [0.523–0.693] |

**Supplementary Table S5.** Paired adaptation changes for the primary clinically anchored models.

| Model | Threshold ΔBA vs strict [II] | No-EB ComBat ΔAUC vs strict [II] | No-EB incremental ΔBA |
|---|---|---|---|
| Clinical (age + CA125) | 0.052 [-0.034–0.091] | N/A | N/A |
| Radiomics + clinical | 0.135 [0.034–0.167] | -0.022 [-0.044–0.009] | -0.047 [-0.141–0.039] |
| DenseNet121 + clinical | 0.193 [0.076–0.193] | -0.007 [-0.026–0.009] | -0.016 [-0.084–0.068] |

| Model | Threshold ΔBA vs strict [II] | No-EB ComBat ΔAUC vs strict [II] | No-EB incremental ΔBA |
|---|---|---|---|
| U-NEXtractor + clinical | 0.070 [-0.047–0.104] | -0.007 [-0.035–0.005] | 0.000 [-0.024–0.045] |
| Fusion U-NEXtractor + clinical | 0.099 [-0.018–0.133] | -0.014 [-0.039–-0.004] | 0.008 [-0.128–0.128] |

*The incremental no-EB ComBat effect compares no-EB ComBat plus threshold adaptation with threshold adaptation alone.*

**Supplementary Table S6.** Prediction stability under no-empirical-Bayes ComBat.

| Model | Spearman ρ [II] | Mean absolute probability change [II] | Locked classification agreement [II] |
|---|---|---|---|
| Radiomics + clinical | 0.638 [0.364–0.825] | 0.160 [0.080–0.280] | 0.658 [0.447–0.843] |
| DenseNet121 + clinical | 0.985 [0.968–0.996] | 0.039 [0.012–0.097] | 0.921 [0.816–1.000] |
| ResNet18 + clinical | 0.995 [0.978–0.999] | 0.055 [0.013–0.157] | 0.974 [0.842–1.000] |
| U-NEXtractor + clinical | 0.999 [0.995–1.000] | 0.010 [0.002–0.035] | 0.947 [0.895–1.000] |
| Fusion DenseNet121 + clinical | 0.495 [0.105–0.739] | 0.130 [0.069–0.217] | 0.737 [0.578–0.842] |
| Fusion ResNet18 + clinical | 0.331 [0.014–0.598] | 0.148 [0.059–0.288] | 0.789 [0.631–0.947] |
| Fusion U-NEXtractor + clinical | 0.974 [0.947–0.988] | 0.020 [0.012–0.040] | 0.947 [0.895–1.000] |

**Supplementary Table S7.** Harmonization coverage and safety status.

| Model | No-EB complete repeats | EB complete repeats | No-EB interpretation | EB interpretation |
|---|---|---|---|---|
| Radiomics + clinical | 200/200 | 0/200 | Included | Excluded |
| DenseNet121 + clinical | 200/200 | 18/200 | Included | Excluded |
| ResNet18 + clinical | 200/200 | 14/200 | Included | Excluded |
| U-NEXtractor + clinical | 200/200 | 25/200 | Included | Excluded |
| Fusion DenseNet121 + clinical | 200/200 | 0/200 | Included | Excluded |
| Fusion ResNet18 + clinical | 200/200 | 0/200 | Included | Excluded |
| Fusion U-NEXtractor + clinical | 200/200 | 0/200 | Included | Excluded |

*Across model, repeat, and held-out-fold attempts, 2,352 empirical-Bayes harmonization failures were recorded; divide-by-zero RuntimeWarnings were the dominant safety-gate trigger.*

**Supplementary Table S8.** Paired strict external discrimination differences versus the clinical baseline.

| Model | ΔAUC (95% CI) | p value | ΔPR AUC (95% CI) | p value |
|---|---|---|---|---|
| DenseNet121 + clinical | 0.136 [-0.030–0.305] | 0.103 | 0.055 [-0.107–0.233] | 0.422 |
| ResNet18 + clinical | 0.063 [-0.154–0.280] | 0.566 | -0.001 [-0.178–0.125] | 0.982 |
| U-NEXtractor + clinical | 0.122 [-0.067–0.303] | 0.193 | 0.031 [-0.145–0.183] | 0.646 |
| Fusion DenseNet121 + clinical | 0.020 [-0.179–0.212] | 0.834 | -0.007 [-0.174–0.119] | 0.900 |
| Fusion ResNet18 + clinical | 0.019 [-0.209–0.243] | 0.858 | -0.013 [-0.190–0.119] | 0.825 |
| Fusion U-NEXtractor + clinical | 0.120 [-0.068–0.301] | 0.199 | 0.031 [-0.145–0.182] | 0.655 |
| Radiomics + clinical | 0.061 [-0.137–0.245] | 0.530 | 0.004 [-0.156–0.132] | 0.959 |

### Supplementary Figures

Supplementary Figure S1. Prediction stability for all imaging representations under no-empirical-Bayes ComBat. Panels show strict-versus-harmonized rank correlation, mean absolute probability change, and locked-classification agreement. Points are medians across 400 held-out fold evaluations; lines are 2.5th–97.5th percentile instability intervals.

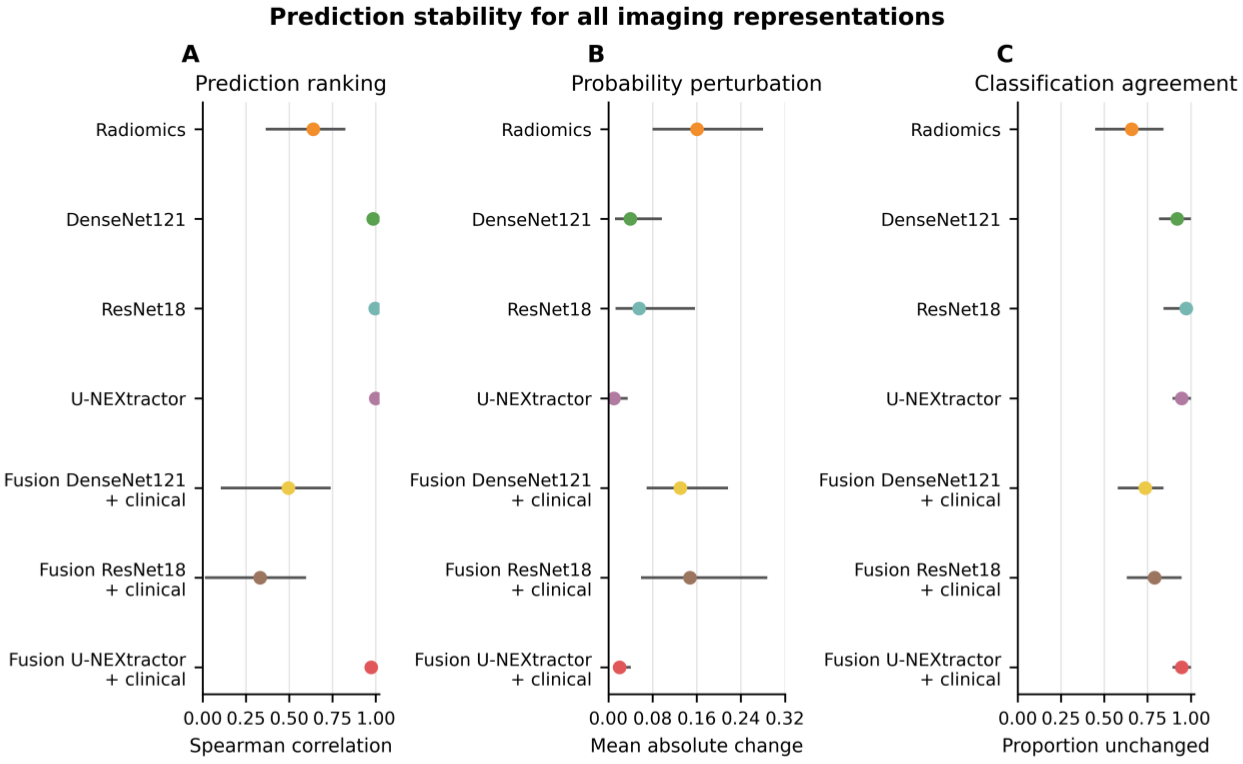

Supplementary Figure S2. Performance across testing conditions for all prespecified models. Heatmaps show fixed supportive internal and strict external estimates and repeated-split medians after threshold adaptation or no-empirical-Bayes ComBat plus threshold adaptation. Clinical-only ComBat cells are not applicable because clinical variables were never harmonized.

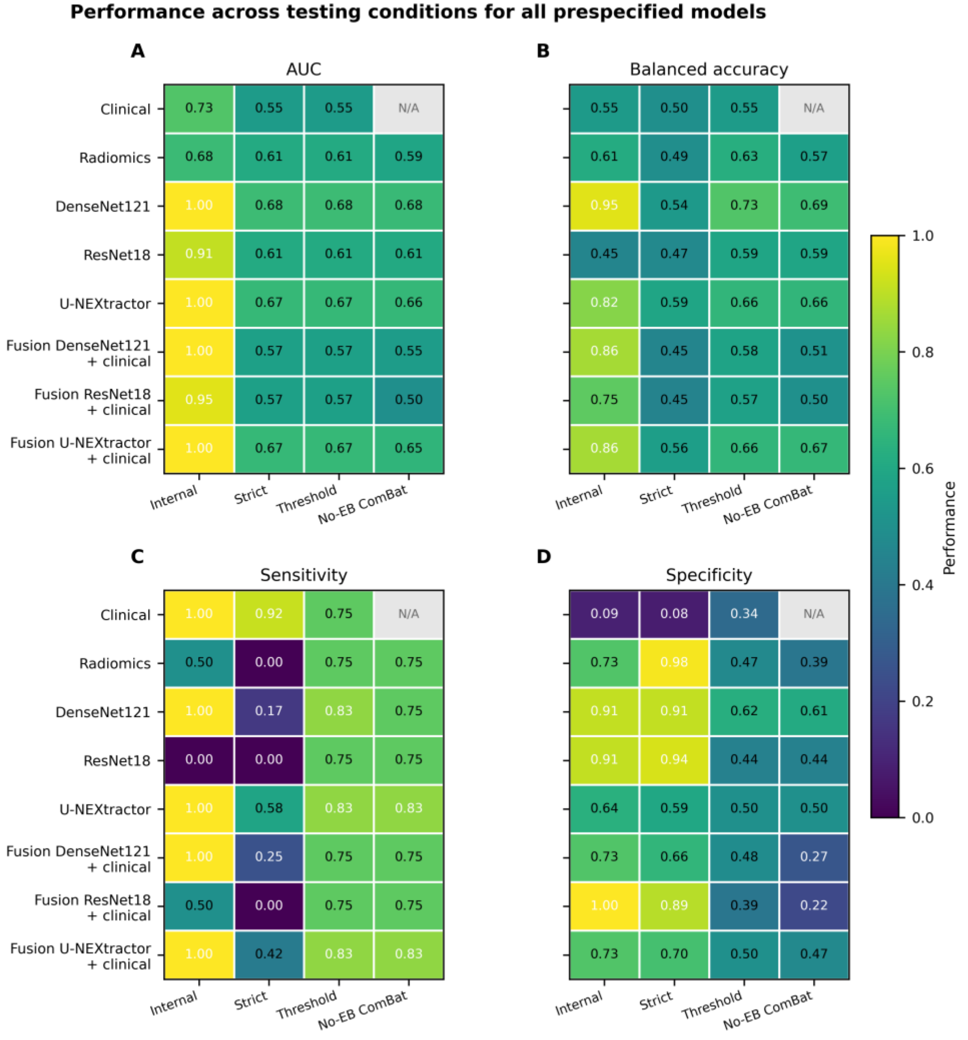


| AUC | Internal | Strict | Threshold | No-EB ComBat |
|---|---|---|---|---|
| Clinical | 0.73 | 0.55 | 0.55 | N/A |
| Radiomics | 0.68 | 0.61 | 0.61 | 0.59 |
| DenseNet121 | 1.00 | 0.68 | 0.68 | 0.68 |
| ResNet18 | 0.91 | 0.61 | 0.61 | 0.61 |
| U-NEXtractor | 1.00 | 0.67 | 0.67 | 0.66 |
| Fusion DenseNet121 + clinical | 1.00 | 0.57 | 0.57 | 0.55 |
| Fusion ResNet18 + clinical | 0.95 | 0.57 | 0.57 | 0.50 |
| Fusion U-NEXtractor + clinical | 1.00 | 0.67 | 0.67 | 0.65 |

| Balanced accuracy | Internal | Strict | Threshold | No-EB ComBat |
|---|---|---|---|---|
| Clinical | 0.55 | 0.50 | 0.55 | N/A |
| Radiomics | 0.61 | 0.49 | 0.63 | 0.57 |
| DenseNet121 | 0.95 | 0.54 | 0.73 | 0.69 |
| ResNet18 | 0.45 | 0.47 | 0.59 | 0.59 |
| U-NEXtractor | 0.82 | 0.59 | 0.66 | 0.66 |
| Fusion DenseNet121 + clinical | 0.86 | 0.45 | 0.58 | 0.51 |
| Fusion ResNet18 + clinical | 0.75 | 0.45 | 0.57 | 0.50 |
| Fusion U-NEXtractor + clinical | 0.86 | 0.56 | 0.66 | 0.67 |

| Sensitivity | Internal | Strict | Threshold | No-EB ComBat |
|---|---|---|---|---|
| Clinical | 1.00 | 0.92 | 0.75 | N/A |
| Radiomics | 0.50 | 0.00 | 0.75 | 0.75 |
| DenseNet121 | 1.00 | 0.17 | 0.83 | 0.75 |
| ResNet18 | 0.00 | 0.00 | 0.75 | 0.75 |
| U-NEXtractor | 1.00 | 0.58 | 0.83 | 0.83 |
| Fusion DenseNet121 + clinical | 1.00 | 0.25 | 0.75 | 0.75 |
| Fusion ResNet18 + clinical | 0.50 | 0.00 | 0.75 | 0.75 |
| Fusion U-NEXtractor + clinical | 1.00 | 0.42 | 0.83 | 0.83 |

| Specificity | Internal | Strict | Threshold | No-EB ComBat |
|---|---|---|---|---|
| Clinical | 0.09 | 0.08 | 0.34 | N/A |
| Radiomics | 0.73 | 0.98 | 0.47 | 0.39 |
| DenseNet121 | 0.91 | 0.91 | 0.62 | 0.61 |
| ResNet18 | 0.91 | 0.94 | 0.44 | 0.44 |
| U-NEXtractor | 0.64 | 0.59 | 0.50 | 0.50 |
| Fusion DenseNet121 + clinical | 0.73 | 0.66 | 0.48 | 0.27 |
| Fusion ResNet18 + clinical | 1.00 | 0.89 | 0.39 | 0.22 |
| Fusion U-NEXtractor + clinical | 0.73 | 0.70 | 0.50 | 0.47 |

Supplementary Figure S3. ComBat completion and safety diagnostics. Panel A shows the proportion of 200 repeats with complete empirical-Bayes harmonization. Panel B shows the distribution of the no-empirical-Bayes harmonized-to-strict median absolute standardized mean-difference ratio. Empirical-Bayes results were excluded from performance interpretation because complete coverage was not achieved.

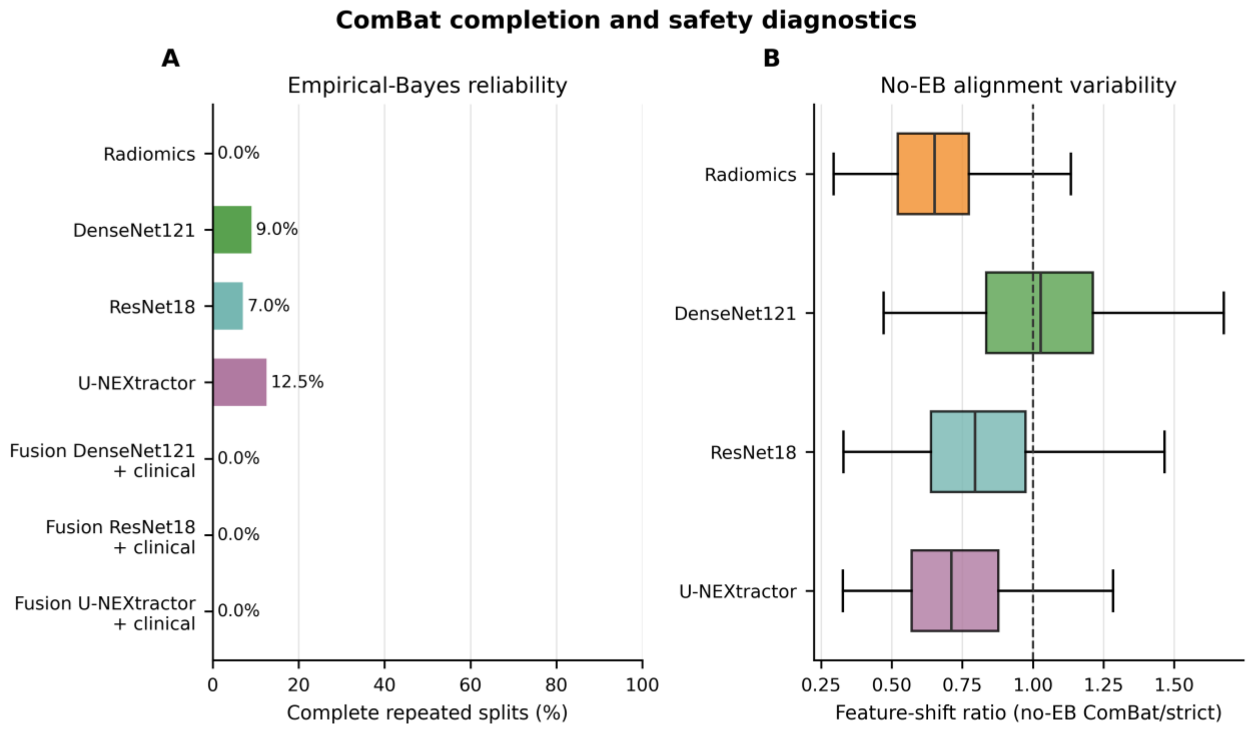


### Abbreviations

ADC = apparent diffusion coefficient; AUC = area under the receiver operating characteristic curve; BA = balanced accuracy; CA125 = cancer antigen 125; CI = confidence interval; DWI = diffusion-weighted imaging; EB = empirical Bayes; FOCUS = reduced field-of-view diffusion-weighted imaging; FN = false negative; FP = false positive; FOV = field of view; II = instability interval; LVSI = lymphovascular space invasion; PR AUC = area under the precision-recall curve; Se = sensitivity; SMD = standardized mean difference; Sp = specificity; TE = echo time; TN = true negative; TP = true positive; TR = repetition time.
